# Model selection reveals the butyrate-producing gut bacterium *Coprococcus eutactus* as predictor for language development in three-year-old rural Ugandan children

**DOI:** 10.1101/2021.03.15.21253665

**Authors:** Remco Kort, Job Schlösser, Alan R. Vazquez, Prudence Atukunda, Grace K.M. Muhoozi, Alex Paul Wacoo, Wilbert F.H. Sybesma, Ane C. Westerberg, Per Ole Iversen, Eric D. Schoen

## Abstract

**Introduction:** The metabolic activity of the gut microbiota plays a pivotal role in the gut-brain axis through the effects of bacterial metabolites on brain function and development. In this study we investigated the association of gut microbiota composition with language development of three-year-old rural Ugandan children.

**Methods:** We studied the language ability in 139 children of 36 months in our controlled maternal education intervention trial to stimulate children’s growth and development. The dataset includes 1170 potential predictors, including anthropometric and cognitive parameters at 24 months, 542 composition parameters of the children’s gut microbiota at 24 months and 621 of these parameters at 36 months. We applied a novel computationally efficient version of the all-subsets regression methodology and identified predictors of language ability of 36-months-old children scored according to the Bayley Scales of Infant and Toddler Development (BSID-III).

**Results:** The best three-term model, selected from more than 266 million models, includes the predictors *Coprococcus eutactus* at 24 months of age, *Bifidobacterium* at 36 months of age, and language development at 24 months. The top 20 four-term models, selected from more than 77 billion models, consistently include *Coprococcus eutactus* abundance at 24 months, while 14 of these models include the other two predictors as well. Mann-Whitney U tests further suggest that the abundance of gut bacteria in language non-impaired children (*n* = 78) differs from that in language impaired children (*n* = 61) at 24 months. While obligate anaerobic butyrate-producers, including *Coprococcus eutactus, Faecalibacterium prausnitzii, Holdemanella biformis, Roseburia hominis* are less abundant, facultative anaerobic bacteria, including *Granulicatella elegans, Escherichia/Shigella* and *Campylobacter coli*, are more abundant in language impaired children. The overall predominance of oxygen tolerant species in the gut microbiota of Ugandan children at the age 24 months, expressed as the Metagenomic Aerotolerant Predominance Index (MAPI), was slightly higher in the language impaired group than in the non-impaired group (*P* = 0.09).

**Conclusions:** Application of the all-subsets regression methodology to microbiota data established a correlation between the relative abundance of the anaerobic butyrate-producing gut bacterium *Coprococcus eutactus* and language development in Ugandan children. We propose that the gut redox potential and the overall bacterial butyrate-producing capacity could be factors of importance as gut microbiota members with a positive correlation to language development are mostly strictly anaerobic butyrate-producers, while microbiota members that correlate negatively, are predominantly oxygen tolerant with a variety of known adverse effects.

## INTRODUCTION

There is an accumulating amount of evidence for a role of the gut microbiota in brain function and development via the so-called microbiota-gut-brain-axis, as recently reviewed by (Cryan et al., 2019). The communication along this axis is bidirectional. Communication from the brain to the gut occurs through signals to change bowel movements and intestinal permeability, which in turn changes the enteric microbiota composition, its metabolic activity and response signal. The gut microbiota signals to the brain via stimulation of intestinal host immune cells, eliciting a cytokine response. In addition, signals are transferred to the brain through bacterial metabolites, including short chain fatty acids. This results in altered neurotransmitter release, hormone secretion and induction of vagus nerve signaling to the brain (Rhee et al., 2009;Bienenstock et al., 2015;Jameson et al., 2020).

In this study we investigated the correlation between gut microbiota composition with the language ability of three-year-old rural Ugandan children, as assessed by the Bayley Scales of Infant and Toddler Development (BSID-III) composite scores for language development (Albers and Grieve, 2007). The scales provide comprehensive development measures for children up to 42 months and have been adapted for appropriate use among children in rural Uganda (Muhoozi et al., 2016). The data used in this study were collected during a follow-up trial of a two-armed, open cluster-randomized education intervention regarding nutrition, child stimulation and hygiene among mothers of children in the Kisoro and Kabale districts of South-West Uganda (Muhoozi et al., 2018). The intervention did not lead to any significant changes in the gut microbiota diversity compared with the control group at phylum or genus level. Neither did we observe any significant differences between the two study groups in the Shannon diversity index at 20-24 and 36 months, respectively. However, the Shannon diversity index of the gut microbiota increased significantly in both study groups from 20-24 to 36 months (Atukunda et al., 2019). Further analysis of the changes associated with the gut microbiota in the transition from 24 months to 36 months revealed that there was a notable shift from autochthonous (endogenous) to allochthonous (plant-derived) *Lactobacillus* species, and a correlation of *Lactobacillus* with stunting, most probably resulting from the change in the children’s diet from breast milk to solid, plant-based foods (Wacoo et al., 2020). As follow-up to this study we further investigate here correlations between the gut microbiota of these children with language development.

It should be noted that predictors for current cognition parameters in children may not only be found in past values of these parameters, but also in current and past gut microbiota compositions. This is supported by longitudinal studies that indicate a maturation program of the human gut microbiome in the first three years of life, consisting of distinct phases of microbiome progression (Backhed et al., 2015;Stewart et al., 2018). Suitable predictors are usually found by fitting models including the predictors being assessed and comparing the fit of the model with the fit of a model that does not include these predictors. This poses a nontrivial problem, because the number of different models that can be fitted grows exponentially with the number of potential predictors, so it is not feasible to fit all possible models and compare their fit. In addition, the predictors can be correlated so that different sets of predictors can explain the response variable of interest equally well. In the present paper, we successfully address the above mentioned problems in data analysis of the gut microbiota from rural Ugandan children. Our key finding is that abundance of butyrate-producing bacterium *Coprococcus eutactus* in the gut microbiota at 24 months predicts language development in these children at 36 months.

## MATERIALS AND METHODS

### Study Design and Data Collection

The data used in this study were collected during a follow-up trial of a two-armed, open cluster-randomized education intervention regarding nutrition, stimulation and hygiene among impoverished mothers of children in the Kisoro and Kabale districts of South-Western Uganda (Muhoozi et al., 2018). The purpose of the study by Muhoozi *et al*. was to assess the effects of a nutrition education intervention, delivered in group meetings to impoverished mothers, on child growth, cognitive development and gut microbiota in rural Uganda. Developmental outcomes were assessed with the Bayley Scales of Infant and Toddler Development (BSID-III) composite scores for cognitive (primary endpoint), language and motor development. Other outcomes included gut microbiota compositions.

Stool samples were collected from 139 children at the age of 20-24 months and at 36 months and shipped to the Netherlands for DNA extraction (Atukunda et al., 2019). Quantitative PCR was performed to determine the relative amount of bacterial template and amplicon sequencing was carried out as previously described (de Boer et al., 2015;Parker et al., 2018). In summary, V4 16S rRNA gene amplicon sequencing was carried out by paired end sequencing conducted on an Illumina MiSeq platform (Illumina, The Netherlands). Taxonomic names were assigned to all sequences using the Ribosomal Database Project (RDP) naïve Bayesian classifier with a confidence threshold of 60% (Wang et al., 2007) and the mothur-formatted version of the RDP training set v.9 (Schloss et al., 2009). All 16S rRNA amplicon paired end reads of the gut microbiota samples sequenced in this study are accessible at BioProject PRJNA517509 (Kort, 2019).

Language development was determined by the Bayley Scales of Infant and Toddler Development 3^rd^ edition (BSID-III) using the language subscale. The BSID-III provides comprehensive development measures with children up to 42 months and has been adapted for appropriate use among children in rural Uganda (Muhoozi et al., 2016;Muhoozi et al., 2018). The BSID-III language component focuses on prelinguistic behaviors, communication and social routines in addition to expressive and receptive language skills. The children’s performance was scored according to the guidelines in the administration manual and the raw scores from expressive and receptive subscales were summed up and converted to composite scores using BSID-III conversion tables. In the reference material of US children the mean score after conversion is 100.

### Model Selection using Mixed Integer Optimization

Model selection strategies should reveal sets of predictors that explain the data equally well, if such is the case. Best subset selection (Miller, 2002) based on Ordinary Least Squares (OLS) returns the best *k* models with *p* predictors each, so that the common predictors in the best models form a solid basis to explain the response variable of interest and the predictors that differ among the best models point to alternative interpretations to explain the same variable. However, until recently, subset selection could only be performed when the total number of predictors *t* is fairly small, say, *t* < 30. Therefore, best subset selection used to be a less attractive model selection technique for research that assesses many parameters. Obviously, one could perform OLS-based forward selection to select predictors (Miller, 2002). This approach has the disadvantage that the resulting models comprise a single path in multidimensional space. That is, there is one model for each number of predictors up to *p*. There is no guarantee that the model with *p* predictors corresponds with the model of the same size from best subset selection.

(Bertsimas et al., 2016) proposed methodology to select the best model with *p* out of *t* predictors with *t* in the 100s. Their approach is based on Mixed Integer Optimization (MIO). The key innovation is that searching unpromising sets of predictors is cut off in an early stage of the calculations so that not all of the models with *p* predictors have to be assessed. In the original form, just one model with *p* predictors is returned along a range of values for *p* extended the original form to obtain the second-best up to *k*-th best models of given size as well (Vazquez et al., 2020). The method thus results in a list of models compatible with the data. The authors further employ a powerful visualization method to reveal possible alternative ways to explain the same variable. For example, one might observe that either the effect of predictor X or the effect of predictor Y is in the best ten models that link language development to four predictors, but the models do not include both of them.

For ease of reference, we call the method of (Vazquez et al., 2020) MIO after its core element. It was developed primarily with applications in statistical design of industrial experiments in mind. The data in these cases usually have few observations and many controllable experimental factors. This is similar to field studies on human microbiota compositions where the number of cases is much smaller than the number of species.

A key element of MIO is best-subset selection, which finds the best fitting model with *p* parameters as measured by the model’s residual sum of squares. Current state-of-the-art algorithms for best-subset selection, as implemented in SAS 9.4 or JMP 14, or in the ‘leaps’ package in R, which is based on (Furnival and Wilson, 1974), do not allow solving the problem when the search is over more than *t*=30 predictors (Vazquez et al., 2020). (Bertsimas et al., 2016) proposed a formulation for the best subset selection in terms of a mixed integer optimization problem. Modern optimization solvers such as (Gurobi, 2017), do permit searching over a large number of potential predictors. The goal function to be minimized is

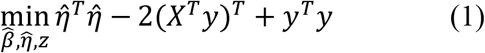

In this equation, 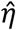is an *N* x 1 vector of fitted values, 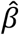is a *t* x 1 vector of coefficients for the regression equation, *y* is the *N* x 1 vector of observations, *X* is an *N* x *t* matrix of predictors, and *z* is a *t* x 1 indicator vector that indicates whether or not the corresponding elements of 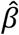are nonzero. The goal function (1) is a version of the residual sum of squares rewritten to reduce the number of quadratic variables from *t* to *N*. This is useful because in our application there are many more potential predictors than there are subjects.

An optimization model allows for the minimization of the goal function under constraints. The constraints proposed by (Bertsimas et al., 2016) are:

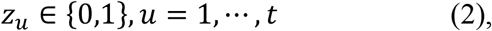

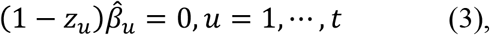

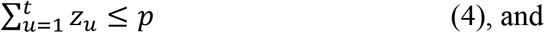

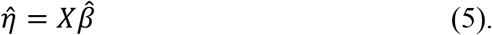

Constraint (2) defines the individual elements *z*_*u*_ of the vector *z* as binary variables. Constraint (3) features the regression coefficients for the individual predictors 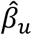. The constraint specifies that 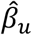 can be nonzero if *z*_*u*_ equals 1 and that 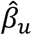 is exactly zero if *z*_*u*_ equals 0. Constraint (4) restricts the regression model to at most *p* nonzero parameters. Finally, constraint (5) defines 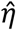as the fitted values matching the coefficients in 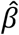. The model (1)-(5) returns for each value of *p* specified by the data analyst the best fitting model as measured by the residual sum of squares. (Vazquez et al., 2020) extended the application potential by proposing further constraints to obtain the second best, third best etc. model for each value of *p*. For example, if parameters 1 and 2 define the best fitting model with *p=2* terms, the constraint z_1_+z_2_ < 2 is added to the constraints (2)-(5) and the model is rerun. The constraints prevent simultaneous inclusion of parameters 1 and 2 in the new model so that a second best model results. (Vazquez et al., 2020) implemented the mixed integer optimization model in Python using (Gurobi, 2017) as the solver of the optimization and used the raster plots of (Wolters and Bingham, 2011) to visualize the models. For this purpose, the predictors are rescaled so that they all have the same length. The raster plot represents each model with *p* parameters as *p* pixels that are darker or lighter according to the size of the respective coefficients. Each predictor has its own horizontal coordinate and each model has its own vertical coordinate. The models are ordered according to the number of nonzero coefficients and, subsequently, their residual sum of squares. Predictors that often occur in the models form a band in the plot.

Promising predictors of the language development of 139 children at 36 months of age were selected for the MIO approach described above. The data included a total of 1170 potential predictors (supplemental file S1), including one parameter indicating whether or not the mother of the child was included in the education intervention group), six anthropometric and cognitive parameters when the children were 24 months 542 gut microbiota composition related parameters at 24 months and 621 parameters at 36 months. Subsequently, the 20 best models were established with 1-4 predictors in terms of their residual standard deviation. The best 4-term models were selected from more than 77 billion models, which is the number of ways one can choose 4 objects out of 1170.

In order to compare the results obtained by the MIO approach to those obtained by a conventional statistical method, the same data were also evaluated by the nonparametric Mann-Whitney U test (Mann and Whitney, 1947). Using this test we investigated which bacterial species had a different abundance in the gut microbiota of children that scored equal or above average for language development when compared with those that scored lower than average. For this purpose, all the 139 children at the age of three years old were divided into a ‘language impaired’ or ‘language below average’ group with a BSD-III score below the mean value of 100 (*n* = 61), and a ‘language non-impaired’ or ‘equal or above average’ group with a BSID-III score of 100 or higher (*n* = 78).

### PCR-based Identification of *Coprococcus eutactus* in Stool Samples

For the experimental identification of *C. eutactus* in stool samples, species-specific primers were designed for the 16S rRNA gene via primer-BLAST™ (Ye et al., 2012): forward-primer 785F 5’-GGGTTCCAAAGGGACTCGG-3’ and reverse primer 1412R 5’-CAGCTCCCTCTTGCGGTT-3’. The oligonucleotides were manufactured by Biolegio™ (Nijmegen, The Netherlands) and delivered in 100 μL TAE-buffer with a concentration of 100 μM. DNA was released from the stool samples in nuclease-free milliQ by heating an Eppendorf tube at 95°C for 10 minutes. The PCR mix contained 12.5 μL GoTaq™ mastermix, 2.5 μL of 10 μM forward primer, 2.5 μL of 10 μM reverse-primer, 5.0 μL nuclease-free milliQ, 2.5 ul template DNA. PCR-samples were placed in the PCR machine (Biometra™, model *Tgradient*) with 1 cycle of 95°C for 5 minutes; 30 cycles of 95°C for 30 seconds, 60°C for 30 seconds and 72°C for 1 minute, completed 72°C for 5 minutes. Products were analyzed by the use of a 1.5% agarose gel with ethidium bromide in TAE-buffer. The PCR was validated by the use of genomic DNA from the cultivated *C. eutactus* type strain ATCC 27759 as a positive control. This strain was obtained from the German Collection of Microorganisms and Cell Cultures (DSM strain number 107541) and cultivated under anaerobic conditions in chopped meat casitone (CMC) medium as described by the supplier.

### The Core Microbiota of Ugandan children at 24 and 36 Months of Age

For the definition of the core, the bacterial 16S rRNA gene amplicon sequencing dataset of the Ugandan children’s feces cohort (139 subjects, measured at 24 months) was used, obtained from the study of (Atukunda et al., 2019). The 1163 bacterial V4-region 16S rRNA gene sequences, delivered in Microsoft™ Excel format, were annotated to bacterial genus and species via the local BLAST in CLC Workbench™ Version 20.0 computational software. All sequence abundances were grouped to species-level and ordered by most prevalent to least prevalent. Bacterial V4-region 16S rRNA amplicon sequences with hits of more than three species with identical identity-scores were grouped to genus level (e.g. *Bifidobacterium*). The core was composed by the top 50 most prevalent bacterial species at a 0.1% relative abundance detection threshold, as described previously (Shetty et al., 2017). From the composed core a heat map was created by MeV™ software (Saeed et al., 2003), thereby including a 0.1 to 100 logarithmic scale for the x-axis threshold at percentage of relative abundance and representing the prevalence via color-scaling for each of the 50 relative abundance detection thresholds.

### Assessment of Metagenomic Aerotolerant Predominance Index (MAPI)

To assess aerobic/anaerobic balance in the gut microbiota samples of our cohort we used the Metagenomic Aerotolerant Predominance Index (MAPI) (Million and Raoult, 2018), based on a previously published database with a list of bacteria and their aerotolerant or obligate anaerobic metabolism (Million et al., 2016). This MAPI index indicates the ratio of the metagenomic relative abundance of aerotolerant species and the relative abundance of strict anaerobes. From the taxonomic assignment of ASV’s of each of the 139 stool bacterial communities of Ugandan children (Supplemental file S1), we calculated the total number of reads that corresponded to strict aerotolerant or anaerobic bacteria. We then calculated the ratio of aerotolerant relative abundance to strict anaerobic relative abundance. This ratio was > 1 for aerotolerant predominance and < 1 for strict anaerobic predominance. In order to fit a lognormal distribution, the natural logarithm of the aerotolerant ratio was calculated for each metagenome for further analysis. The metagenomics aerotolerant predominance index (MAPI) corresponds to the variable ‘ln(Ae/Ana)’.

### Ethical Approval

All mothers gave written or thumb-printed, informed consent to participate and could decline an interview or assessment at any time. The study was approved by The AIDS Support Organisation Research Ethics Committee (no. TASOREC/06/15-UG-REC-009) and by the Uganda National Council for Science and Technology (no. UNCST HS 1809) as well as by the Norwegian Regional Committee for Medical and Health Research Ethics (no. 2013/1833). The trial was registered at clinicaltrials.gov (NCT02098031).

## RESULTS

### Abundance of *Coprococcus eutactus* is a predictor for language development

The application of the MIO approach to identify predictors for language development in Ugandan children at 36 months of age resulted in the raster plot of Figure 1A. For this visualization, we normalized the predictors and the language development score such that their means are zero and their standard deviations are one. A coefficient thus expresses the increase in the response, in terms of multiples of its standard deviation, if a predictor is increased by one standard deviation. As co-occurrences can only be recorded in models with two or more parameters, we ignore models with a single parameter in our evaluation. The red vertical band with horizontal axis label 5 shows that, with a few exceptions, the best models with 2-4 parameters include the language development of the children at 24 months and that its coefficient is positive. This parameter is included in 52 of the 60 models with 2-4 parameters. The figure shows that in 7 of the 8 remaining cases, cognition at 24 months (horizontal axis label 4) replaces language development at 24 months in the model. The MIO methodology shows here alternative explanations of the same data by correlated predictors. Indeed, the Pearson correlation coefficient of the language ability and cognition parameters equals 0.7. In spite of this correlation, the much higher frequency of occurrence of the language ability at 24 months suggests that this parameter should be included in favor of cognition at 24 months. Further red bands can be observed at horizontal axis labels 281 and 563, respectively. These bands correspond with relative abundances of *Coprococcus eutactus*, and *Bifidobacterium* from the gut microbiota at 24 months and 36 months of age, respectively. The abundance of *C. eutactus* occurs in 42 of the 60 models with 2-4 parameters, while the abundance of *Bifidobacterium* occurs in 19 of these models. A total of 18 of the models include both parameters. Species identities were verified with the BLAST tool. They led to a species assignment on the basis of a 100% identity match with the partial 16S rRNA sequence of *Coprococcus eutactus* strain ATCC 27759 (Holdeman and Moore, 1974) over the total length of the sequenced V4 region of 253 base pairs. The assignment of the species *Coprococcus eutactus* is unambiguous, but sequences of *Bifidobacterium longum* and *Bifidobacterium breve* are both aligning to the 16S rRNA V4 sequence with a match of 100% identity, therefore we refer to parameter 563 as a match to the *Bifidobacterium longum* group (see also below).

**Figure 1.**
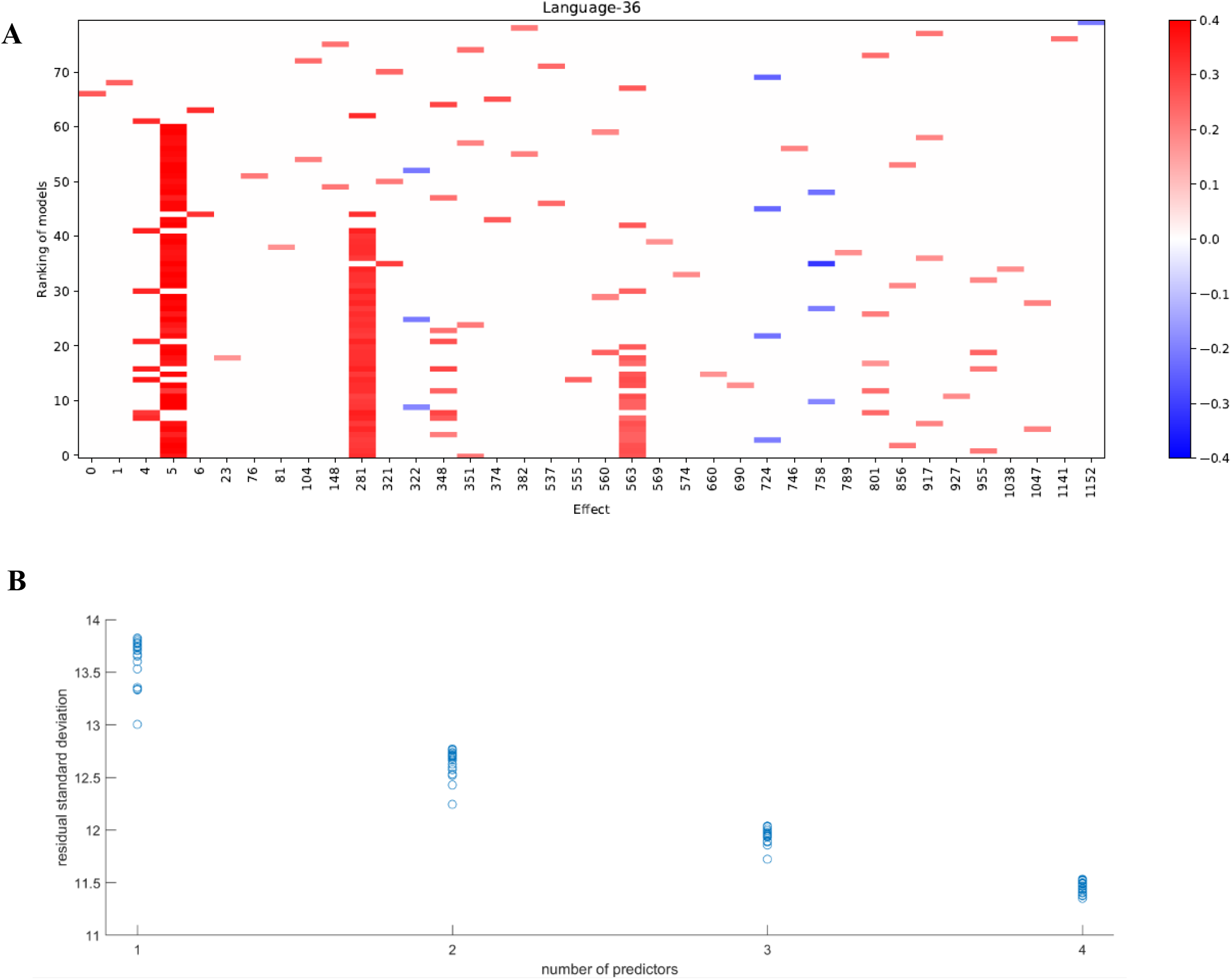
The 20 best fitting models of 1 up to 4 terms linking language development of children aged 36 months to membership of the intervention or control group, values of 6 developmental parameters of the same children when aged 20-24 months, their microbiota composition at that age, and microbiota composition at 36 months. **A)** Predictors in the models. Language development and all predictors are normalized to a mean of 0 and a standard deviation of 1. Horizontal labels correspond with the predictors’ identification number in the data file. The parameters include the following scores at 24 months: the intervention indicator (0); height to age, HAZ (1); weight to age, WAZ (2); weight to height, WHZ (3); cognition (4); language development (5); motoric development (6). These are followed by the gut microbiota parameters at 24 months (parameters 7-548) and at 36 months (parameters 548-1169). Vertical coordinates 1-20, 21-40, 41-60 and 61-80 show best fitting models with 4, 3, 2 and 1 terms, respectively. Blue pixels correspond with negative coefficients and red pixels correspond with positive coefficients. Intensity of the pixels increases with size of the model coefficients. **B)** Residual standard deviations plotted against the number of terms in the models for language development; language development in original units.

Figure 1A further shows that some predictors enter the fitted models occasionally. The most frequently occurring parameter after the *Bifidobacterium longum* group is *Intestinibacter bartlettii* (previously known as *Clostridium bartlettii*) at 24 months of age (horizontal label 348). This identification was based on a unique 100% identity match of the V4 amplicon with the partial 16S rRNA gene sequence of the type strain *Intestinibacter bartlettii* strain WAL 16138 (Song et al., 2004). As this parameter enters only in 8 out of the 60 models, there is no powerful evidence that it should be included in a regression model. The residual standard deviations for the 80 models in Figure 1A were plotted against the number of predictors in the models (Figure 1B). The figure shows that, for four predictors, many of the 20 subsets explain the data equally well, so this application of a method to reveal the common elements in these subsets is warranted. Exploration of the common elements points to a 3-parameter model with parameters 5 (language development of the children at 24 months), 281 (abundance of *C. eutactus*) and 563 (abundance of the *B. longum* group at 36 months), respectively. This model turns out to be the best 3-parameter model. The figure shows that its residual standard deviation clearly stands out from the remaining 19 models. We conclude that a model including these 3 parameters explains the data best.

The linear regression model for BSID-III language development at 36 months with all parameters on their original scale is summarized in Table 1. For the intercept and each predictor in the model, the entries in the last four columns show the regression coefficient, its standard error, the ratio of the coefficient to its standard error (t Ratio) and the *P*-value of this ratio. The residual standard deviation of the model is 11.7 based om 135 degrees of freedom. This measure quantifies the variability unexplained by the model. The model’s F value equals 21.5. This measure indicates how much larger the model mean square is when compared to the unexplained variance. Finally, the adjusted R^2^ value of 31% is the percentage of variation accounted for by the model, adjusted for the number of parameters.

**Table 1.**
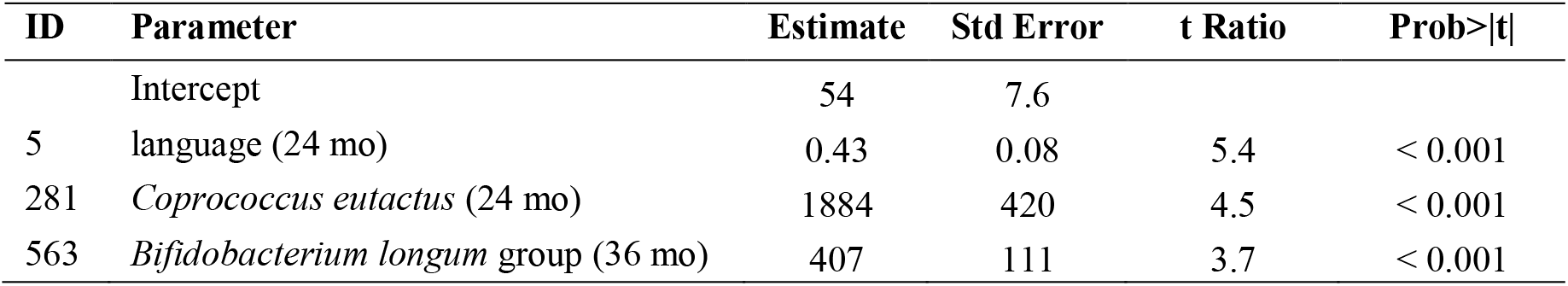
Coefficients for the prediction model for language development. Coefficients were calculated for the BSID-III scores of children aged 36 months by the MIO approach as indicated in Figure 1. ID, identification number in data file; Std Error, standard error; Prob >| t|, *P*-value. Residual standard deviation = 11.7; degrees of freedom = 135; model F value = 21.5; Adjusted R^2^ (%) = 31.

| ID | Parameter | Estimate | Std Error | t Ratio | Prob>\|t\| |
| --- | --- | --- | --- | --- | --- |
|  | Intercept | 54 | 7.6 |  |  |
| 5 | language (24 mo) | 0.43 | 0.08 | 5.4 | < 0.001 |
| 281 | *Coprococcus eutactus* (24 mo) | 1884 | 420 | 4.5 | < 0.001 |
| 563 | *Bifidobacterium longum* group (36 mo) | 407 | 111 | 3.7 | < 0.001 |

All the model coefficients are positive so that higher values of previous BSID-III language development, previous abundance of *C. eutactus* and current abundance of *B. longum* point to higher language development at 36 months.

The large values of the abundance coefficients can be explained by the measurement scale. The observed relative abundances of *C. eutactus* at 24 months (ID 281) and *B. longum* at 36 months of age (ID 563) are at most 4.5% (see Figure 2 for scatter interval plots of the relative abundances of *C. eutactus* and *B. longum*). The model in Table 1 includes the language development score recorded when the children were at the age of 24 months. The interpretation of this finding is that children that had the same development score when they were of that age, differ in their subsequent language development according to their microbiota composition at that age and the composition at their current age.

**Figure 2.**
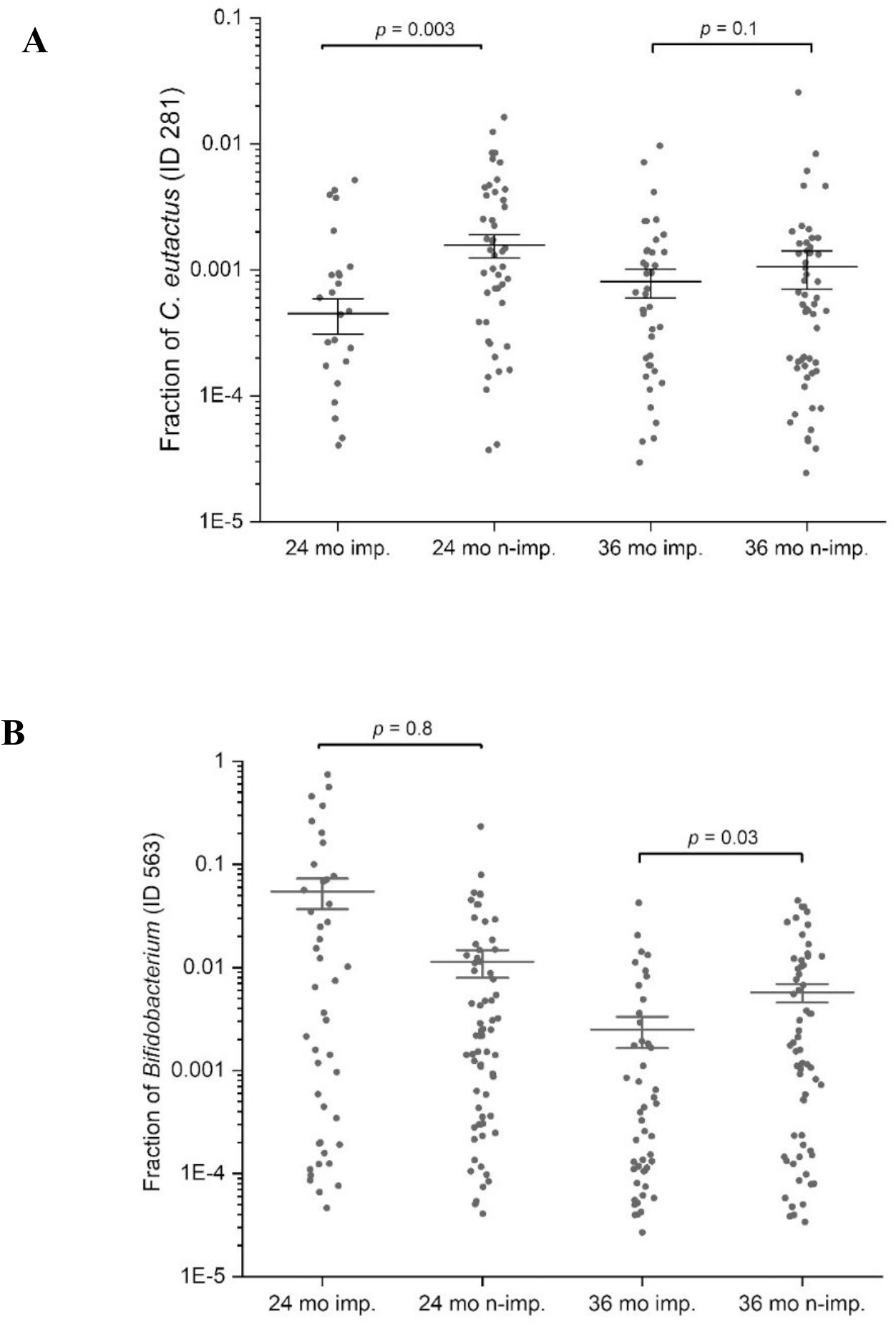
Scatter interval plots of the fraction or relative abundance of *C. eutactus* and *Bifidobacterium*. **A)** Fraction of *Coprococcus eutactus* (amplicon sequence variant ID 281) in the gut microbiota of 139 Ugandan children at the age of 24 and 36 months, **B)** Fraction of *Bifidobacterium longum* group (amplicon sequence variant ID 563) in the gut microbiota of Uganda children aged 24 and 36 months. *P*-values were calculated with the two sided Mann-Whitney U test for language impaired (*n* = 61) and language non-impaired groups (*n* = 78) of the children.

### Increased prevalence of *Coprococcus eutactus* in the core gut microbiota over time

In order to evaluate the prevalence of *C. eutactus* in the microbiota of children of 24 months and 36 months in relation to other highly abundant members of the intestinal microbiota, we carried out a comparative analysis of the core gut microbiota from the Ugandan children at 24 and 36 months (Figure 3). It should be noted that for this analysis all ASV’s assigned to the same species have been pooled together. Both cores appear to be rather comparable in composition (80% of the species are present in both cores). However, at 24 months only the top three species, including *Faecalibacterium prausnitzi Prevotella copri*, and *Blautia wexlerea*, were highly prevalent (> 90%, detection threshold 0.1%), while a set of ten species is highly prevalent among Ugandan children at 36 months, in line with a decrease in the variation in the gut microbiota composition among children at higher age. The overall prevalence and abundance of *Bifidobacterium* species increased at 36 months compared to 24 months (from position 24 to 10), although this is not the case for ASV’s matching to *Bifidobacterium longum* (see ASV ID 563, Figure 2B), in agreement with the notion that the relative abundance of this species reduces when children are no longer breast fed. The butyrate producing species *Faecalibacterium prausnitzii* was the most prevalent bacterial species in the datasets of both ages and is present in all Ugandan children in our study at the age of 36 months. Noteworthy, both microbiota cores clearly show a typical *Prevotella* gut microbiota type, in agreement with previous observations that led to the assignment of *Bacteroides* and *Prevotella* as biomarkers of diet and lifestyle in Western and non-Western subjects, respectively (Gorvitovskaia et al., 2016), and references herein. Accordingly, *Prevotella* species, such as *P. copri*, show much higher relative abundance among the majority of subjects in both heat maps of 24 and 36 months than *Bacteroides* species, represented in the core only by *B. xylanolyticus*. The gut bacterium *C. eutactus* is also represented in both cores, be it at relatively low prevalence and abundance levels; species position 44 at the age of 24 months and position 37 at the age of 36 months. The prevalence of the *C. eutactus* ASV ID 281 among the Ugandan children in this study increased from 24 months to 36 months from 62% to 81%, although the average relative abundance was slightly lower (Figure 2A). This increased prevalence over time was also evident in the core gut microbiota; all 11 ASV’s matching to *C. eutactus* showed an increase from 38% to 44% at 0.1% abundance threshold (Figure 3). These observations and our best fitting model are in agreement with the notion that early acquirement of *C. eutactus* (before or at 24 months) is a beneficial factor for language development.

**Figure 3.**
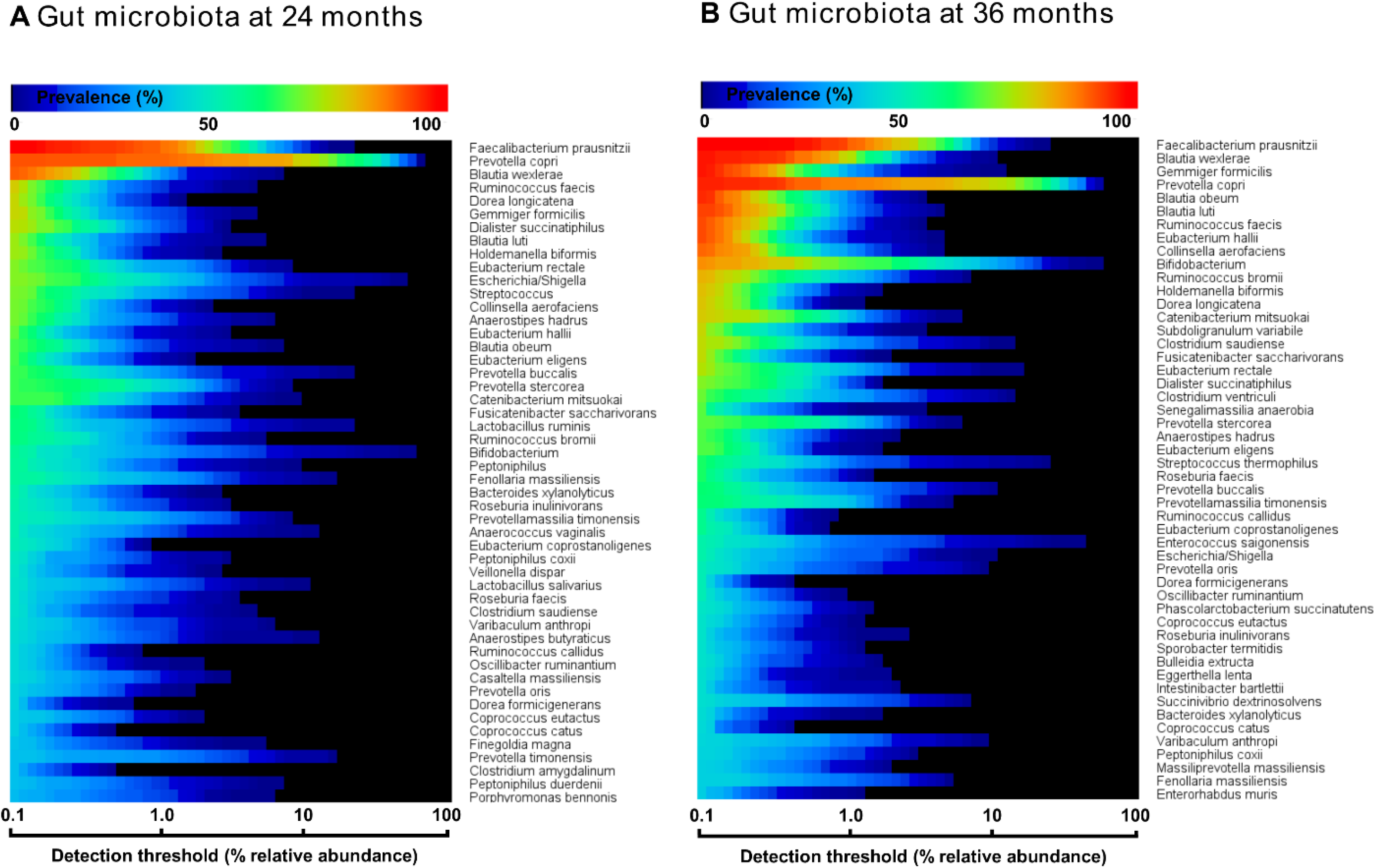
Heat maps of top 50 most prevalent bacteria in the fecal microbiota of Ugandan children at the age of 24 months (**A**) and 36 months (**B**), as determined by 16S rRNA gene amplicon sequencing. The color gradient indicates the prevalence (see top-legend) at the detection threshold of the relative abundance (%) presented at the x-axis with a logarithmic scale. The y-axis indicates the order of most prevalent bacteria at a detection threshold of 0.1% abundance. Unambiguous species assignments include *Dialister succinatiphilu, D. propionicifaciens*; *Lactobacillus salivarius, L. ruminis*; *Clostridium saudiense, C. disporicum*: *Varibaculum anthropi, V. cambriense*; *Prevotella oris, P. albensis, P. salivae*; *Clostridium amygdalinum, C. methoxynbenzovorans*.

### Butyrate-producing species more abundant in children with above average language development

The presence of the butyrate-producing bacterium *C. eutactus* was confirmed in the fecal samples of the Ugandan children in our cohort by PCR using specific primers designed in this study for *C. eutactus*. PCR-analysis of a random set of stool samples from the rural Ugandan children showed a product in 75% of the fecal samples in line with the prevalence range (62% to 81%) found for *C. eutactus* in our 16S rRNA gene sequence data from the children’s stool samples. In order to further substantiate the results obtained by the MIO approach, we also checked with a conventional statistical method (the Mann-Whitney U test) which bacterial species had a different abundance in children that scored equal or above average for language development when compared with children that scored below average. We first checked for the presence of the specific amplicon sequence variants (ASV’s) predicting language development by the MIO approach. We found that the relative abundances of the identified ASV’s in our best fitting model of *C. eutactus* at 24 months (ID 281) and *Bifidobacterium* at 36 months (ID 563) were significantly different in both groups according to the two sided Mann-Whitney U test, with *P*-values of 0.003 and 0.03, as presented in Figure 2.

Out of the 542 gut microbiota ASV parameters at 24 months, 397 matched to a bacterial species with an identity score of 97% or higher. Using the latter composition parameters, we employed the two sided Mann-Whitney U tests to explore on a per species basis differences in abundance of these parameters between three-year old children that scored equal or above average for language development and children that scored below average. If these two language groups would not differ in microbiota composition, we would expect that 20 out of the 397 tests have *P*-values below 0.05. Instead, twenty-five of these tests had such a *P*-value. Table 2 lists the corresponding ASV’s. Nineteen ASV’s were more abundant in the equal or above average group and six ASV’s were more present in the below average group. Among these were also other unique sequences matching to parameters identified in the predictive models as presented in Figure 1, including *Bifidobacterium* (ID 23), and *Intestinibacter bartlettii* (ID 348).

**Table 2.** Two-tailed Mann-Whitney U test for relative abundance of bacterial species equal or above average and below average language ability groups. A two-tailed test was performed for bacterial relative abundances of the gut microbiota of Ugandan children at the age of 24 months for language impaired (*n* = 61; BSID-III scores < 100) and non-impaired groups (*n* = 78; BSID-III scores ≥ 100). Bacterial species (based on BLAST searches of amplicon sequence variants, ASV’s) listed in the table had a *P*-value below 0.05. Species with identity scores below 97% were excluded from the list in the table. Unambiguously assigned bacterial species are indicated by superscripts. The ASV-match of *Bifidobacterium catenulatum*^1^ is identical to that of *B. pseudocatenulatum, B. kashiwanohense, B. tsurumiense, B. callitrichidarum* and *B. gallicum* (assigned to the *catenalatum* group); *Bifidobacterium adolescentis*^2^ is identical to that of *B. faecale* and *B. stercoris* (assigned to the *adolescentis* group); *Clostridium disporicum*^3^ identical to *C. saudiense*; *Bifidobacterium longum*^4^ to *B. breve* (assigned to the *longum* group).

| ID | Bacterial species | Identity (%) | P-value | core member | obligate anaerobic |
| --- | --- | --- | --- | --- | --- |
| **Species with higher relative abundance in non-impaired language group** |  |  |  |  |  |
| 23 | *Bifidobacterium catenulatum*<sup>1</sup> | 99.6 | 0.003 | yes | yes |
| 20 | *Bifidobacterium adolescentis*<sup>2</sup> | 99.6 | 0.005 | yes | yes |
| 357 | *Faecalibacterium prausnitzii* | 97.6 | 0.006 | yes | yes |
| 14 | *Bifidobacterium adolescentis*<sup>2</sup> | 100.0 | 0.008 | yes | yes |
| 406 | *Holdemanella biformis* | 97.6 | 0.010 | yes | yes |
| 294 | *Roseburia hominis* | 100.0 | 0.010 | no | yes |
| 316 | *Eubacterium eligens* | 99.2 | 0.013 | yes | yes |
| 513 | *Campylobacter troglodytis* | 98.8 | 0.015 | no | no |
| 25 | *Bifidobacterium adolescentis*<sup>2</sup> | 99.6 | 0.020 | yes | yes |
| 466 | *Faecalibacterium prausnitzii* | 97.6 | 0.021 | yes | yes |
| 78 | *Prevotella copri* | 99.6 | 0.022 | yes | yes |
| 348 | *Intestinibacter bartlettii* | 100.0 | 0.024 | no | yes |
| 352 | *Terrisporobacter petrolearius* | 99.6 | 0.032 | no | yes |
| 318 | *Bacteroides xylanolyticus* | 98.0 | 0.035 | yes | yes |
| 390 | *Clostridium disporicum*<sup>3</sup> | 97.2 | 0.040 | yes | yes |
| 280 | *Coprococcus eutactus* | 99.6 | 0.042 | yes | yes |
| 399 | *Catenibacterium mitsuokai* | 97.6 | 0.044 | yes | yes |
| 519 | *Campylobacter troglodytis* | 98.4 | 0.047 | no | no |
| **Species with higher relative abundance in impaired language group** |  |  |  |  |  |
| 219 | *Granulicatella elegans* | 100.0 | 0.0005 | no | no |
| 57 | *Parabacteroides* | 97.2 | 0.015 | no | yes |
| 31 | *Bifidobacterium longum*<sup>4</sup> | 99.6 | 0.027 | yes | no |
| 529 | *Escherichia/Shigella* | 99.6 | 0.028 | yes | no |
| 528 | *Escherichia/Shigella* | 100.0 | 0.034 | yes | no |
| 521 | *Campylobacter coli* | 97.2 | 0.041 | no | no |

A number of other striking features emerge from this Mann-Whitney U test. The list in Table 2 contains five unique 16S rRNA gene amplicon sequence variants which show a match with the genus *Bifidobacterium*. Although the V4 16S rRNA gene amplicon sequence does not allow for unambiguous assignment of species for this genus, we can assign these ASV’s to three distinct *Bifidobacterium* species groups: the *Bifidobacterium catenelatum, adolescentis* and *longum* groups (see Table 2). Members of the first two groups show at 24 months a positive correlation with language development. Relative abundance of members of the *longum* group show at 24 months a negative correlation with language development, but a positive correlation at 36 months (Figure 1; Figure 2; Table 2). At this point it is not clear why the relative abundance of the *B. longum* species group at 24 months, known to be beneficial and dominant in infants, correlate negatively with language development, in contrast to species from *B. catenulatum* and *adolescentis* groups, which are generally more prevalent in adults (Arboleya et al., 2016). Many of the bacterial species listed in Table 2, which are more abundant in the above average group, are known butyrate producers, including *Coprococcus eutactus, Faecalibacterium prausnitzii, Holdemanella biformis, Roseburia hominis, Clostridium disporicum* and *Catenibacterium mutsuokai* (Vital et al., 2014). The SCFA butyrate has been implicated to play a role in brain function, as further discussed below.

### Increased predominance of oxygen tolerant species in children impaired in language development

While 17 out of 19 ASV’s with significant scores in the above average group matched with strictly anaerobic species (all except for *Campylobacter troglodytis*), we identified only one ASV matching with a strictly anaerobic bacterium (*Parabacteroides*) among the significant scores in the below average group (Table 2). This result is in line with the notion that a relatively high redox potential in the environment of the gut is an adverse condition for language development. It should be noted that majority of the ASV’s in the below average group (4 out of 6) match to species with known adverse effects in humans, including *Granulicatella elegans* (Table 2). Although known to be part of the normal intestinal human microbiota, this species has often been implicated in adverse conditions. In addition, aerobic *Escherichia/Shigella*, and *Campylobacter coli* species are known as major foodborne pathogens, causing the widely occurring diseases shigellosis and campylobacteriosis, which lead to severe diarrhea, in particular at relatively high prevalence among children in the developing world. A box plot of the MAPI indices among all the children in both language development groups indicated a slight difference (*P* = 0.09) between the two groups (Figure 4).

**Figure 4.**
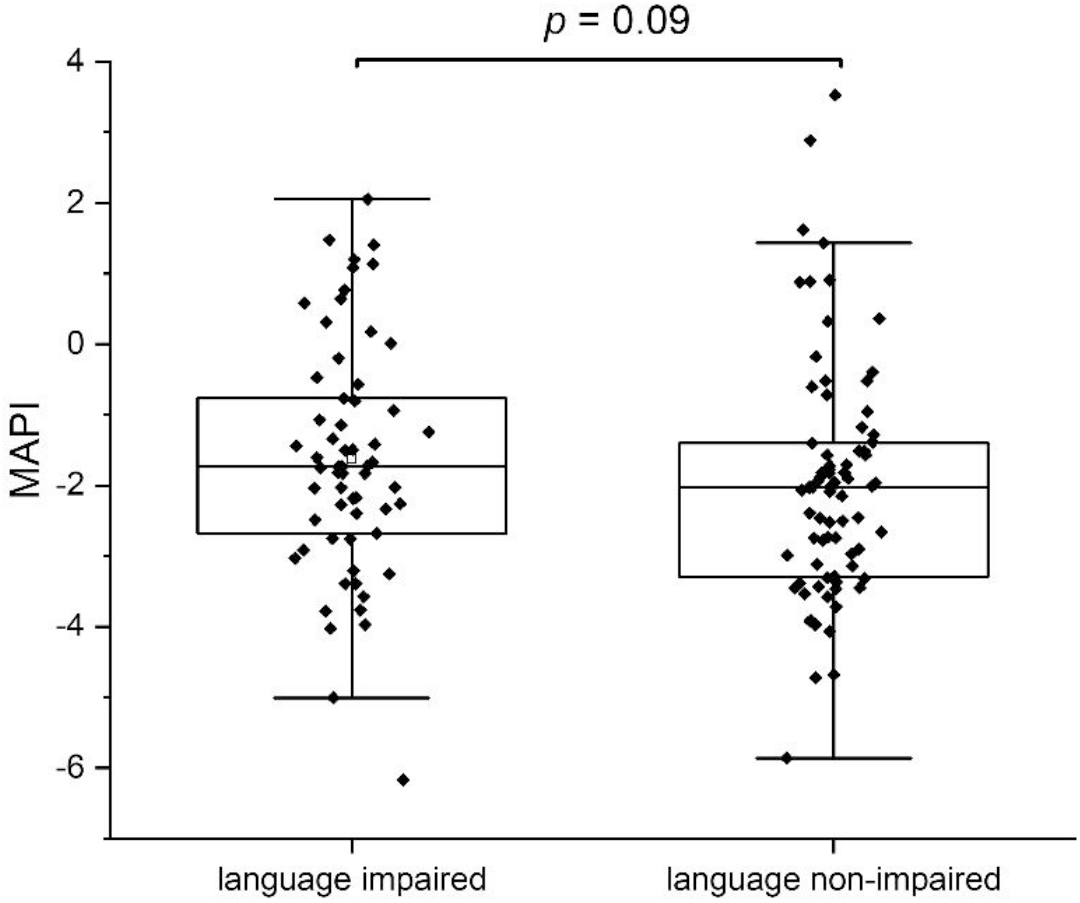
The Metagenomic Aerotolerant Predominance Index (MAPI). The index is presented in box plots for the groups of language impaired (*n* = 61) and language non-impaired children (*n* = 78).

## DISCUSSION

### The value of alternative prediction models

For the analysis presented in this paper, we identified promising predictors of language development in a field study from a large set of potential predictors that are likely to be correlated. Because of this correlation, it was imperative to use methods that could reveal alternative explanations of the same data. In the field data we studied, we indeed found substantial correlations among the observed microbiota abundances that could serve as potential predictors (4164 pairwise correlations were larger than 0.5).

Revealing alternative explanations of the same data requires the fitting of multiple models. We used MIO in our model search. The strong point of this approach is that one can impose constraints relevant for the data at hand. We used this option in our ranking of the second best down to 20^th^ best models. Of particular use were the models with 3 and 4 predictors. There was a clear best 3-predictor model among 20 alternative models. This model included the language ability of the children at 24 months, the abundance of *Coprococcus eutactus* in microbiota taken at 24 months, and the abundance of *Bifidobacterium longum* in microbiota taken at 36 months.

The fact that this model is clearly better than the alternatives suggests that we should include the 3 predictors mentioned in any case. However, there might still be additional predictors that could improve the model fit. This was investigated by fitting 4-parameter models as well.

There was no clear best 4-parameter model. However, *C. eutactus* abundance at 24 months was consistently present in all 4-parameter models, while the other two predictors in the best 3-parameter model were included in 14 of the 20 best 4-parameter models. By focusing on the common predictors present in the best models, we believe that we avoided overfitting the data. The remaining predictors were present in at most 5 out of the 20 best 4-predictor models. We conclude that there is no clear evidence favoring inclusion of a fourth predictor.

A further use of constraints in the MIO approach can help finding good models that include synergistic or antagonistic effects of the microbiota species. However, MIO is still limited in the size of the models it can handle. In particular, it is computationally infeasible to arrive at the best 5-term model based on 1170 potential model terms. As there are 1163 individual predictors involving microbiota composition, synergistic or antagonistic effects among the species would increase this number with 0.5 ×1163 × (1163 - 1) = 675,703 further terms. It is infeasible to have a successful model search among this number of terms.

### Importance of early-life acquirement of the butyrate-producing *Coprococcus eutactus* for language development

One of the most intriguing findings of this work is the correlation between the abundance of members of saccharolytic clostridia in the gut of Uganda children at 24 months with the composite score for language development of the children at 36 months. We identified *Coprococcus eutactus* (42 out of 60 models) and *Intestinibacter bartlettii* (8 out of 60 models). They belong to the Lachnospiraceae and Peptostreptococcaceae, respectively, both families within the Clostridia, a class of obligatory anaerobic spore-forming bacteria. Both species produce short chain fatty acids (SCFA’s), the primary end-products of fermentation of non-digestible carbohydrates that become available to the gut microbiota and gut epithelial cells. The SCFA’s are mainly produced through saccharolytic fermentation of carbohydrates. While *C. eutactus* is known to produce the SCFA’s formate, acetate and butyrate (Holdeman and Moore, 1974), *Intestinibacter bartlettii* produces the SCFA’s isobutyrate and isovalerate (Song et al., 2004). It is well established that SCFA’s, in particular butyrate, are important substrates for maintaining the colonic epithelium, elicit effects on lipid metabolism and adipose tissue at several levels, in appetite regulation and energy intake, and play a role in regulation of the immune system (Morrison and Preston, 2016). In addition, butyrate has been shown to protect the brain and enhance plasticity in animal models for neurological disease. In agreement with a role for the production of butyrate in the gut for improved language development, studies with animal models show that butyrate is able to reverse stress-induced decrease of neurotrophic factors and cognition impairment both at early and later stages of life (Valvassori et al., 2014). A number of mechanisms have been attributed to the beneficial role of butyrate in brain function, including its action as a histone deacetylase inhibitor and as an activator of G protein-coupled receptors (GPR’s); a lower level of histone acetylation is a characteristic of many neurodegenerative diseases, and butyrate has been shown to activate GPR109a, potentially leading to anti-inflammatory effects in the brain (Bourassa et al., 2016).

The most consistent predictor in our MIO models for language development at 36 months was the abundance of *Coprococcus eutactus* in gut microbiota when the children were 24 months of age. This is in agreement with the concept of a maturation program with distinct phases of microbiota compositions, where earlier phases can affect health outcomes later in life (Backhed et al., 2015;Stewart et al., 2018). The dynamics of the relative abundance of *C. eutactus* was highlighted in a study on the human infant gut microbiome in development and in progression towards type 1 diabetes (Kostic et al., 2015). This longitudinal study indicated a maximum of *C. eutactus* relative abundance in healthy infants at approximately 24 months, while the abundance of *C. eutactus* type 1 diabetes predisposed children remained at constant, at relatively low levels in the first years of life. So far we only have analyzed the gut microbiota in children at 24 and 36 months in our cohort, thus at this moment we cannot yet make any substantiated statements about the longitudinal development of the gut microbiota in our cohort. However, the results in our study are in agreement with a model that holds that relatively high levels of *C. eutactus* at 24 months are beneficial, as they are present in the group of children with above average language development at 36 months.

A number of other uncertainties and limitations should be considered in the interpretation of our results. Among all hypervariable regions of 16S rRNA gene, the V4 region used in this study ranks first in sensitivity as a marker for bacterial and phylogenetic analysis (Yang et al., 2016). Nevertheless, these amplicon sequence libraries allow in some cases only a classification of microbiota members on the genus level. Therefore, we carefully examined all assignments to the species level in this study. Overall, the correlation between genomes of closely related species suggests that it may be effective to predict functions encoded in an organism’s genome. A recent study showed phylogeny and function to be sufficiently linked that prediction of function from 16S rRNA gene amplicons can provide useful insights (Langille et al., 2013). However, in our view metagenome sequencing to reveal the full genetic capacity of the gut microbiota, intervention studies with *C. eutactus* in a germ-free mouse model and *in vivo* metabolite measurements are required to acquire additional evidence on a beneficial role of butyrate production and additional neuroactive potential of the gut bacterium *C. eutactus* in cognitive development.

### Relative abundance of *Coprococcus eutactus* correlates to multiple cognitive outcomes

Interestingly, a recent study on the neuroactive potential of the gut microbiota with a large cohort (Flemish Gut Flora Project; n = 1,054) revealed that butyrate-producing *Coprococcus* bacteria were consistently associated with higher quality of life indicators and depleted in depression (Valles-Colomer et al., 2019). The authors of this study performed a module-based examination of metabolic pathways by members of the gut microbiota in order to investigate its neuroactive potential. They observed that a gene encoding for the synthesis of 3,4-dihydroxyphenylacetic acid (a metabolite of the neurotransmitter dopamine) was strongly associated with the presence of *C. eutactus* and quality of life indicators. Notably, a second metabolic module, which co-varied with quality of life indicators in their cohort, is the synthesis of isovalerate. This ability to synthesize this SCFA happens to be present in *Intestinibacterium bartlettii* (Song et al., 2004), which is the species matching to ASV ID 348 in our best fitting models.

A further evaluation of the current scientific literature confirms that the relative abundance of the genus *Coprococcus*, and in particular the species *C. eutactus*, correlates with other cognitive outcomes. A lower relative abundance of *Coprococcus* was found in autistic patients compared to neurotypical controls (Table 3). An independent study confirmed lower levels of fecal acetic acid and butyrate in autistic subjects (Liu et al., 2019). A decreased relative abundance of *C. eutactus* was also observed in fecal samples and mucosal biopts from Russian and American patients with Parkinson’s disease (PD), respectively (Table 3). In both studies, potentially anti-inflammatory, butyrate-producing genera, *Coprococcus, Faecalibacterium* and *Blautia* were significantly more abundant in feces of controls than PD patients, feeding the hypothesis that an altered gut microbiota could contribute to inflammation-induced development of PD pathology (Keshavarzian et al., 2015). A cross-sectional study on schizophrenia patients also indicated that the level of butyrate producing bacterial genera, including *Coprococcus, Blautia* and *Roseburia* significantly decreased in comparison to healthy controls. The observed differences in microbiota compositions were proposed as a basis for the development of microbiota-based diagnosis for schizophrenia (Shen et al., 2018). However, it is clear that among these differences, *i*.*e*. a decrease of a number of butyrate-producing bacterial genera, a similar correlation can be observed for very different adverse cognitive outcomes, including the impaired language development with Ugandan children in our study.

**Table 3.** Correlations between *Coprococcus eutactus* and human mental health outcomes. The table includes information about cohort, sample size, statistical test, *P*-value and study reference.

| Genus species | Finding | Cohort | Sample size (*n*) | *P*-value | Statistical test | Study reference |
| --- | --- | --- | --- | --- | --- | --- |
| *Coprococcus* | Depleted in cohort participants with depression | Flemish Gut Flora project | 1054 | < 0.05 | Covariance test | (Valles-Colomer et al., 2019) |
| *Coprococcus* | Depleted in cohort participants with depression | Dutch lifeline DEEP | 1063 | < 0.05 | Covariance test | (Valles-Colomer et al., 2019) |
| *Coprococcus* | Lower relative abundance in autistic patients compared to neurotypical controls | American children (20 neurotypical and 20 autistic) | 40 | 0.001 | Mann-Whitney U test | (Kang et al., 2013) |
| *Coprococcus* | Lower relative abundance in Parkinson's-diseased patients compared to healthy controls | American adults (34 Parkinson's patients and 31 healthy controls) | 65 | 0.03 | Kruskall-Wallis test | (Keshavarzian et al., 2015) |
| *Coprococcus eutactus* | Lower relative abundance in Parkinson-diseased patients compared to healthy controls | Siberian adults (89 Parkinson's patients and 66 healthy controls) | 157 | 0.03 | White's *t*-test | (Petrov et al., 2017) |
| *Coprococcus* | Relative abundance reduced in schizophrenia patients | 64 schizophrenia patients and 53 healthy controls | 117 | 0.004 | Principal coordinate analysis<br>Welch's *t*-test | (Shen et al., 2018) |
| *Coprococcus eutactus* | Predictor in gut microbiota at 24 months for language development at 36 months | Rural Ugandan children | 139 | < 0.001 | All subsets regression | This study |

We looked in this study for other overall differences between bacterial gut communities in the language impaired and language non-impaired groups of children in our study and found higher levels of oxygen tolerant species in the first group. This finding concerns specific, potentially pathogenic species with significant higher relative abundance in the language impaired group (*Granulicatella elegans, Escherichia/Shigella, Campylobacter coli*), but also to slight differences in the overall MAPI index. As this index indicates an aerotolerant predominance for MAPI > 0 and anaerobic predominance for MAPI < 0, it is clear that both groups have an anaerobic predominance of bacterial species in the gut. Apparently, the increase of a number of oxygen tolerant species in the language impaired group is not so much reflected by the overall MAPI index. Possibly, this results from the fact that the Ugandan children in our study group are not severely malnourished, as they are on average moderately stunted (−3 < HAZ < −2). More severe malnourishment could have led to the overall depletion of anaerobic bacteria and proliferation of oxygen tolerant bacteria, as shown in the gut microbiota of severely malnourished children (Million et al., 2016). In order to confirm the findings in this study, we propose to repeat the analysis and investigate cognitive development as a function of the MAPI index in a similar cohort. In parallel, we propose to set up an intervention study aiming at the reduction of the gut redox potential as a stimulus to create a better growth environment for beneficial, strictly anaerobic gut bacteria, including *Coprococcus eutactus* and other butyrate producers identified in this study.

## Data Availability

All data in this manuscript is available in the supplemental file S1 and accessible at BioProject PRJNA517509

https://www.ncbi.nlm.nih.gov/bioproject/517509

## Author Contributions Statement

EDS developed the models, interpreted MIO outcomes, and made Table 1. RK and EDS drafted the manuscript. ARV carried out the MIO analysis and made Figure 1. RK made figures 2, 3 and 4. GM and PA carried out the field work in Uganda, collected and analyzed the data of the education intervention study. APW supported in microbiota data analysis. ACW and POI designed the education intervention study and analyzed developmental data. JS carried out the PCR, Mann Whitney U test, analysis for the core microbiota, and made Tables 2 and 3. WS supported in the translation of the results. All authors reviewed and approved the final manuscript.

